# An Effective Automated Algorithm to Isolate Patient Speech from Conversations with Clinicians

**DOI:** 10.1101/2022.11.29.22282914

**Authors:** Theo Jaquenoud, Sam Keene, Neveen Shlayan, Alex Federman, Gaurav Pandey

## Abstract

A growing number of algorithms are being developed to automatically identify disorders or disease biomarkers from digitally recorded audio of patient speech. An important step in these analyses is to identify and isolate the patient’s speech from that of other speakers or noise that are captured in a recording. However, current algorithms, such as diarization, only label the identified speech segments in terms of non-specific speakers, and do not identify the specific speaker of each segment, e.g., clinician and patient. In this paper, we present a novel algorithm that not only performs diarization on clinical audio, but also identifies the patient among the speakers in the recording and returns an audio file containing only the patient’s speech. Our algorithm first uses pretrained diarization algorithms to separate the input audio into different tracks according to nonspecific speaker labels. Next, in a novel step not conducted in other diarization tools, the algorithm uses the average loudness (quantified as power) of each audio track to identify the patient, and return the audio track containing only their speech. Using a practical expert-based evaluation methodology and a large dataset of clinical audio recordings, we found that the best implementation of our algorithm achieved near-perfect accuracy on two validation sets. Thus, our algorithm can be used for effectively identifying and isolating patient speech, which can be used in downstream expert and/or data-driven analyses.

## I. Introduction

Speech has long served as a rich source of information for clinicians to evaluate and diagnose patients [1], [2]. Most commonly, the content and cadence of speech can help experts assess patients’ mental and cognitive health [3]–[5]. With recent advances in machine learning (ML) algorithms [6], [7], it is becoming possible to automatically identify disorders or disease biomarkers from digitally recorded patient speech [8]. An important step in achieving this goal is to identify and isolate the patient’s speech from other speech or noise that are captured in a recording of their conversation with a clinician.

This identification process is generally referred to as speaker *diarization*, which is the task of automatically segmenting and labeling an audio recording containing speech according to the identity of the speakers (Figure 1). Accomplishing this task entails simultaneously addressing the challenges of identifying when a person is speaking and recognizing who the speaker is. Diarization has a number of useful applications, particularly as part of larger workflows when combined with other speech processing techniques, such as an automatic video captioning system [9], [10].

**Fig. 1.**
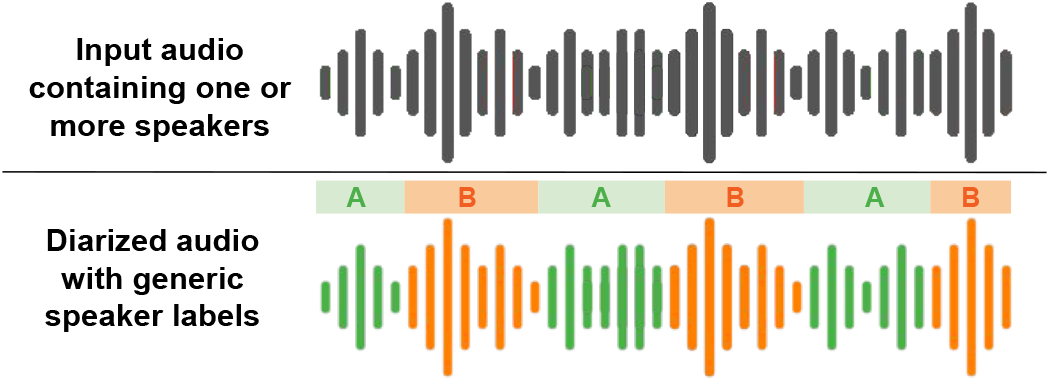
Standard input and output of a diarization algorithm. The input consists of any digital audio recording of speech with one or more speakers. The output is a labeling (e.g., speaker A or B) indicating who, if anyone, is speaking at each moment in the recording.

In clinical research, diarization is often used to identify patient speech that can be input to ML models capable of diagnosing certain conditions, or providing biomarkers for cognitive or mental health. Weiner et al. developed a diarization algorithm as part of a workflow for the automatic screening of dementia [11]. Hanai et al. developed diarization methods for recorded neuropsychological exams [12], which were used by Lin et al. in a longitudinal study to identify digital voice biomarkers for cognitive health [13]. Garoufis et al. used diarization methods as a part of speech analysis algorithms to detect relapses in patients with psychotic disorders such as bipolar disorder and schizophrenia [14]. Hansen et al. investigated the role of diarization in developing generalized speech-based emotion recognition models, particularly with the goal of identifying depression and remission of the condition [15].

While studies like the above have established diarization as an important step in clinical studies involving automated speech analysis (ASA), this step itself is rarely a central focus of these studies. Instead, researchers often implement or develop their diarization algorithms specifically for their workflow and to fit the pathology of interest to them. As a result, there are no standalone patient speech diarization algorithms for clinical applications. Perhaps more importantly, current diarization algorithms label the identified segments only in terms of different non-specific speakers, such as speaker A and B (Figure 1), but do not identify the specific speaker of each segment, e.g., clinician and patient. This nonspecific labeling is not sufficient for a clinical setting, where the patient’s, clinician’s and others’ speech often needs to be considered separately, since they convey different information. This suggests that the field of clinical ASA would greatly benefit from having a standalone tool for identifying and isolating speech only from a patient during the course of a clinical encounter. The output of such a tool could then be used in downstream analyses for identifying and monitoring various conditions, such as dementia, Parkinson’s disease, and affective and psychotic disorders [3]–[5].

In this paper, we present a novel algorithm that not only performs diarization on clinical audio, but also identifies the patient among the speakers in the room and returns an audio file containing only the patient’s speech. We evaluated this method on a large, novel dataset of audio recordings of primary care visits.

## II. Proposed Method

Here, we describe the audio dataset used, as well as our patient speech identification and isolation algorithm.

### A. Audio dataset

The audio dataset used in this paper originated from an NIH-funded study to develop methods for automated detection of mild cognitive impairment (MCI) in primary care practices for adults aged 50 years and older. Patients were recruited in order to form a representative sample of Mount Sinai Health System patients within the target age group. Their demographic information is detailed in Table I. Following approval from the Mount Sinai Institutional Review Board (IRB) and informed consent from all of the participants, high-definition audio was recorded for 854 complete conversations during primary care visits between patients and their clinicians. To the best of our knowledge, this is one of the largest clinical audio datasets collected for research purposes [8].

**TABLE I.** Demographic information of patient cohort

|  |  | Total<br>(n=854) |
| --- | --- | --- |
| Age | Mean (Std. Dev.)<br>(in years) | 67.39 (8.13) |
| Sex | Female | 477 (55.9%) |
|  | Male | 377 (44.1%) |
| Ethnicity | White | 257 (30.1%) |
|  | African American | 279 (32.7%) |
|  | Asian | 18 (2.1%) |
|  | Hispanic | 215 (25.1%) |
|  | Other/Unknown | 85 (10.0%) |

The conversations were recorded using a Tascam DR-10L portable audio recorder with a lavalier (clip-on) microphone attached to the patient. This placement of the recorder on or near the target speaker is a standard setting for collecting patient audio in clinical research studies [17]. While most audio was recorded in high-resolution with a sample rate of 48kHz, the files were down-sampled to 16kHz prior to any additional processing to reduce computation time. No other filtering or denoising was performed. Due to the nature of the visit, the recordings were generally long, with a duration averaging 30min:04s. They were also unstructured, meaning the conversations did not follow specific question-answer formats or prompts like in several other studies [11], [12], [15].

The overall purpose for which this audio dataset was collected was to automatically identify patients with cognitive impairment from their recordings. As such, the first step of this identification was to isolate the patient’s speech from the long, unstructured recording, for which the proposed algorithm (Figure 2) was developed. This algorithm is described in the following subsections.

**Fig. 2.**
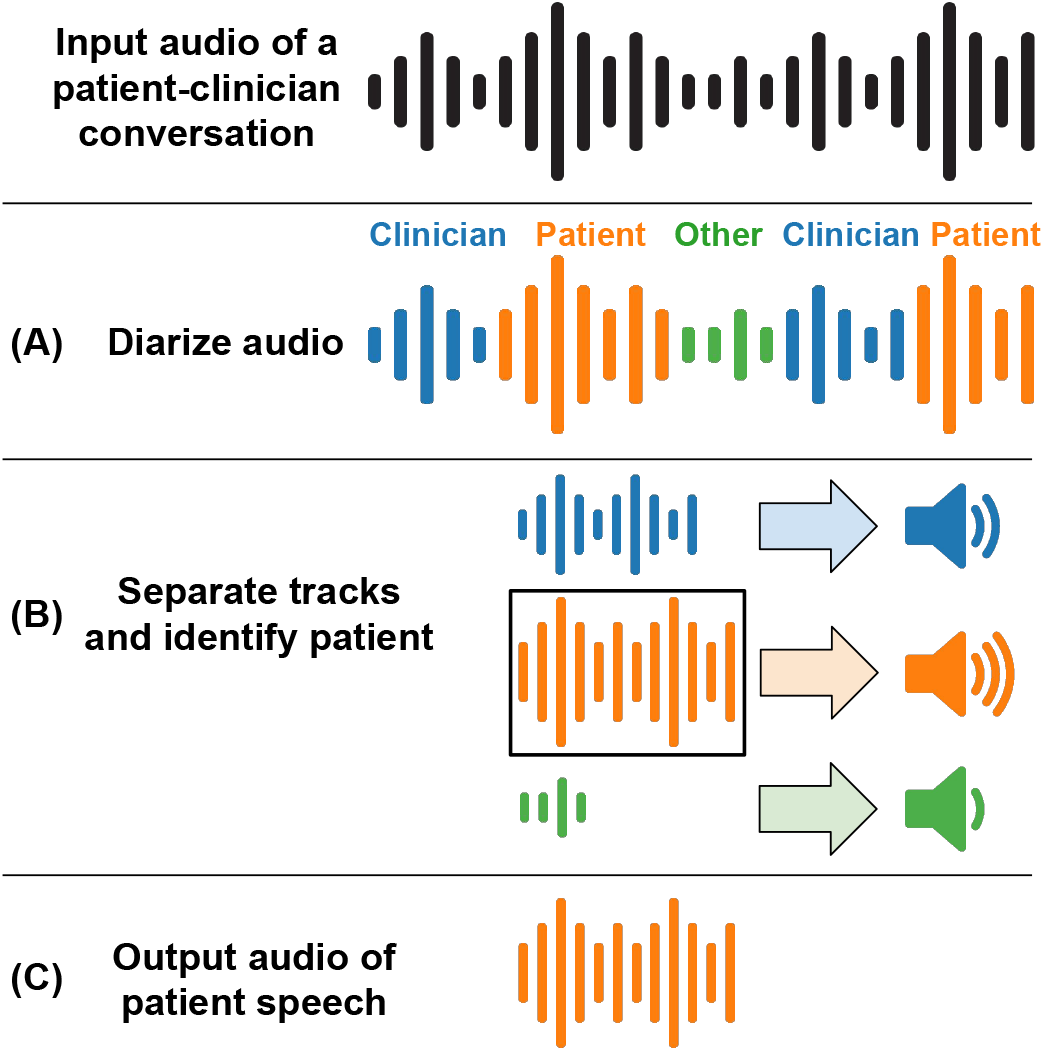
Our proposed workflow for patient speech identification and isolation from audio recordings of patient-clinician conversations. (A) First, the pretrained pyannote-audio diarization algorithm [16] is used to annotate the audio file in terms of generic speakers (i.e., Patient, Clinician and Others, although these specific roles have not been determined at this stage). (B) Next, the audio file is separated into tracks according to the speaker label. The average power of each audio track is then used to determine which speaker is the patient. (C) Finally, the patient’s audio track is returned.

### B. Speaker diarization

The pretrained model in the widely used open-source pyannote-audio toolkit [16] was used for the first step of our algorithm (Figure 2(A)), i.e., general diarization. This model was chosen due to its good performance on a range of diarization datasets, including that of the third DIHARD speech diarization challenge [18], which contains clinical dialog.

This model performs the diarization task in five steps: feature extraction, speech activity detection, speaker change detection, speaker representation and clustering. The features extracted are a combination of standard speech audio features and those inferred from neural networks. These vectors are used as input for the next three steps, producing a set of segments containing speech from generic speakers. The set of segments is then clustered, such that each cluster corresponds to a generic speaker (e.g., A and B in Figure 1). Given an audio file, the model returns a dictionary of segments. Each segment contains a pair of start and end times, a unique segment identifier, and a generic label for the corresponding identified speaker (Figure 1).

After diarization, our algorithm iterated over the unique speakers and used the start and end times of their identified segments to copy content from the source audio file into corresponding audio buffers. Each speaker’s buffer was then analyzed separately (Figure 2(B)).

### C. Isolation of patient speech

Before implementing the final, novel patient speech isolation step of our algorithm (Figure 2(B,C)), we first visualized the labeled sequences resulting from pyannote-audio’s diarization as a color-coded timeline, as illustrated by the three examples in Figure 3. Sequences (i) and (ii) exemplify effective diarization, while sequence (iii) reveals some flaws of diarization models that may affect our algorithm. Given the nature of the patient-clinician conversation in our dataset, we could formulate this step of our algorithm on the assumption that there should be only two speakers. However, examples like Figure 3 (iii) illustrated that one speaker could sometimes be mistaken as two or more speakers. This observation motivated several quantitative methods for identifying the patient among the labeled two or more speakers in the diarization.

**Fig. 3.**
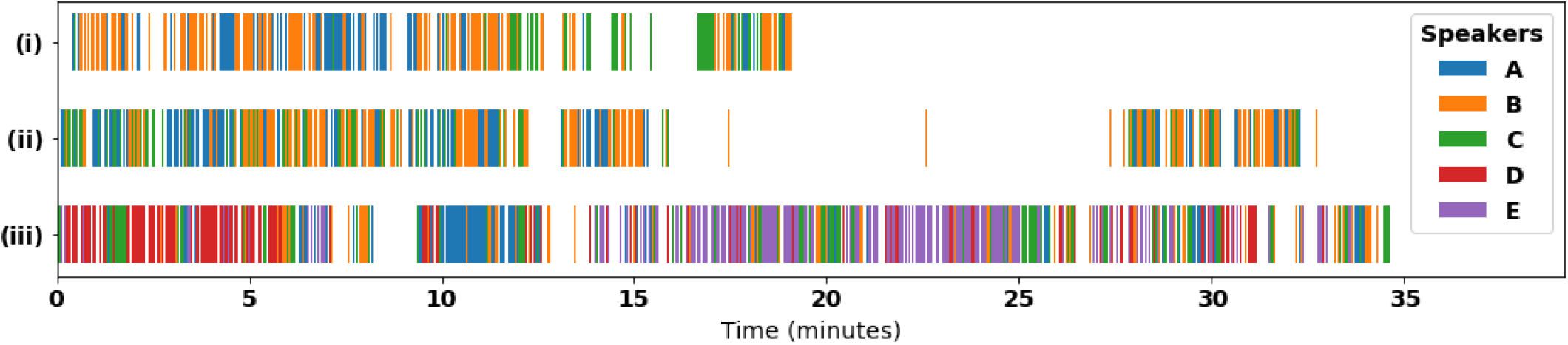
Three examples of color-coded, speaker-labeled timelines for recordings in our dataset diarized with pyannote-audio: (i) and (ii) show typical examples of apparently successful diarizations, in which predominantly two speakers A and B are identified, and a minority of the recordings are attributed to a third speaker C. This matches our *a priori* knowledge of the conversation being recorded (i.e. between a patient and a physician), while (iii) shows an erroneous example where a larger portion of the recording is misclassified as a number of other speakers (C-E).

Our methods relied on the fact that the target speaker, i.e., the patient, was wearing the lavalier microphone that recorded the audio, and was thus expected to sound louder than the clinician and other speakers. While not universal, such recordings are commonplace in clinical research settings [8], [17].

To quantify the loudness of each speaker, the average power of the signal in their corresponding audio buffer was calculated as follows. Digital audio is recorded as a discrete series of data points, or samples, each representing the volume of the recorded sound at a particular instant. Given an audio signal *x* indexed by instants *i*, and of length *N*, we can compute the average power by taking the sum of the absolute squares of the samples *x(i)* and dividing it by *N* (Equation (1)) [19].

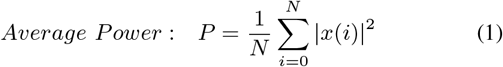

This average power metric was then used to help identify the patient from within the generically labeled speakers using the following two methods:

leftmargin=*

- **Method 1:** Identify the loudest speaker, i.e., the buffer with the highest average power, as the patient.
- **Method 2:** Identify the patient as the louder of the two most prominent speakers, i.e., consider only the two longest buffers and select the one with the higher average power.

Method 1 relies only on the assumption that the patient was the loudest speaker, while Method 2 makes the additional assumption that there were only two speakers in the recording, namely the patient and the clinician. Once the patient was identified by either method, their corresponding audio buffer was produced as the output of our overall algorithm.

### D. Implementation Details

All aforementioned software was written and executed in Python 3.7 [20]. The diarization model in II-B was implemented in PyTorch [21], and can be adapted to run on either a central processing unit (CPU) or a graphics processing unit (GPU). While the model is faster on a GPU, the CPU implementation was used to enable greater parallelization. The algorithms were run on Oracle cloud infrastructure compute instances. While different configurations were leveraged in different executions of the algorithm, as per the availability in the cloud setup, a representative execution used the Oracle Linux 8.5 operating system running 32 third-generation AMD cores with a base clock speed of 2.55Ghz and 512 GB of memory. All patient privacy constraints were fully respected in all the computations as per IRB and HIPAA requirements.

## III. Experimental Results and Analysis

Below, we describe the results of the application of the proposed patient speech identification and isolation algorithm to our dataset of audio recordings of patient-clinician conversations.

### A. Evaluation methodology

The main goal of our evaluation was to assess how well our algorithm was able to isolate only the patient’s speech from the audio recording. However, a rigorous evaluation of this task would have required manually labeling the entire recording in terms of the speaker at each instant. In addition to this labeling being very labor-intensive, it also raises concerns about preserving patient privacy. For these reasons, a comprehensive evaluation was outside the scope of this study.

Thus, we used an alternative, more practical methodology to assess the performance of the proposed algorithm, i.e., the combination of the diarization and two patient identification methods described in Section II. For this evaluation, three non-overlapping 30 second segments were randomly sampled from each of the diarized and isolated audio files, representing more than 25% of the length of an average file after processing. A rater then determined if the majority of each sampled segment contained speech from the patient, allowing for overlapping voices, which would indicate that the patient had been accurately identified and isolated for that recording. If needed, the rater used unprocessed source recording to get additional context to identify the patient. The performance of the algorithm for the full patient speech file was then considered correct if all three of the segments were determined to be accurately isolated by the rate. This evaluation was repeated for all isolated patient audio files in the validation set under consideration, and the accuracy of the proposed algorithm was calculated as in Equation (2).

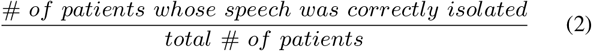

### B. Quantitative and qualitative evaluation

We first applied the above evaluation methodology to a validation set consisting of 20 randomly sampled patient-clinician recordings from our entire audio dataset. Two raters separately applied this evaluation methodology to the outputs of our algorithm for this validation set. One rater was the lead author of this paper. The other was a clinical research coordinator who played a role in recruiting patients and recording audio of their visits with clinicians. Both the raters briefly reviewed a sufficient number of these recordings to be able to reliably identify patient speech in the randomly selected audio segments. However, this knowledge of theirs had no bearing on the actual operation of our diarization and isolation algorithm, and was only leveraged in this evaluation.

The specific goal of this evaluation was to compare the performance of Methods 1 and 2 described in Section II-C for the patient speech isolation task. The raters determined that Method 1 performed accurately for 100% of reviewed outputs, while Method 2 did so for only 75.0% of the cases. The assessments of the raters were fully consistent, i.e., the inter-rater agreement was 100%.

To further confirm the above results, the lead author of this paper repeated this evaluation with a randomly sampled validation set containing an additional 40 patient-clinician recordings. For this set, Method 1 performed with 97.5% accuracy, while Method 2 achieved 77.5%. These results confirmed that Method 1 was more appropriate for our task of the identification and isolation of patient speech, and was used for the last step in the final implementation of our proposed algorithm (Figure 2(C)).

We also qualitatively analyzed potential reasons for the differential performances of Methods 1 and 2. In particular, we observed that Method 2, which selected only the louder of the two most prominent speakers, was generally not successful in cases exemplified by Figure 3(iii), where the pyannote-audio diarization model identified many more than two speakers. If either the patient’s or the clinician’s speech was mistakenly fragmented into multiple speakers, it was often the case that neither of the two most prominent speakers were the patient. In such a case, Method 2 was not able to select the correct audio buffer as the patient’s speech, while Method 1 still could.

A similar concern was possible with Method 1. Specifically, it was possible that, by simply choosing the loudest speaker regardless of how often or prominently they appeared in the audio, we might sometimes misidentify a short portion of loud audio as the patient. For example, if a nurse briefly walked into the primary care visit room and spoke loudly, that short segment of audio may have the highest average power (loudness) despite not being from the patient. However, we observed no occurrences of this problem in our validation sets, contributing to Method 1’s strong performance, and further supporting our choice of this method for the patient speech isolation step (Figure 2(C)).

### C. Data compression ability of patient speech isolation

In addition to accurately isolating patient speech which researchers can focus on for downstream analysis, our algorithm can also enable several computational savings, as we observed with our implementation and platform (Section II-D). For instance, the algorithm is able to eliminate long silences (e.g., in Figure 3(ii)) and other such artifacts in the original recordings. This leads to a substantial shortening of the resultant audio recordings, as demonstrated for our entire audio dataset in Figure 4. This has implications for storage and memory efficiency, as the storage requirement of our dataset was reduced by more than 80% from about 148 GB to about 26 GB. This data compression can enable a more judicious use of high-performance computing resources by sophisticated downstream ML and other data-driven analyses [22].

**Fig. 4.**
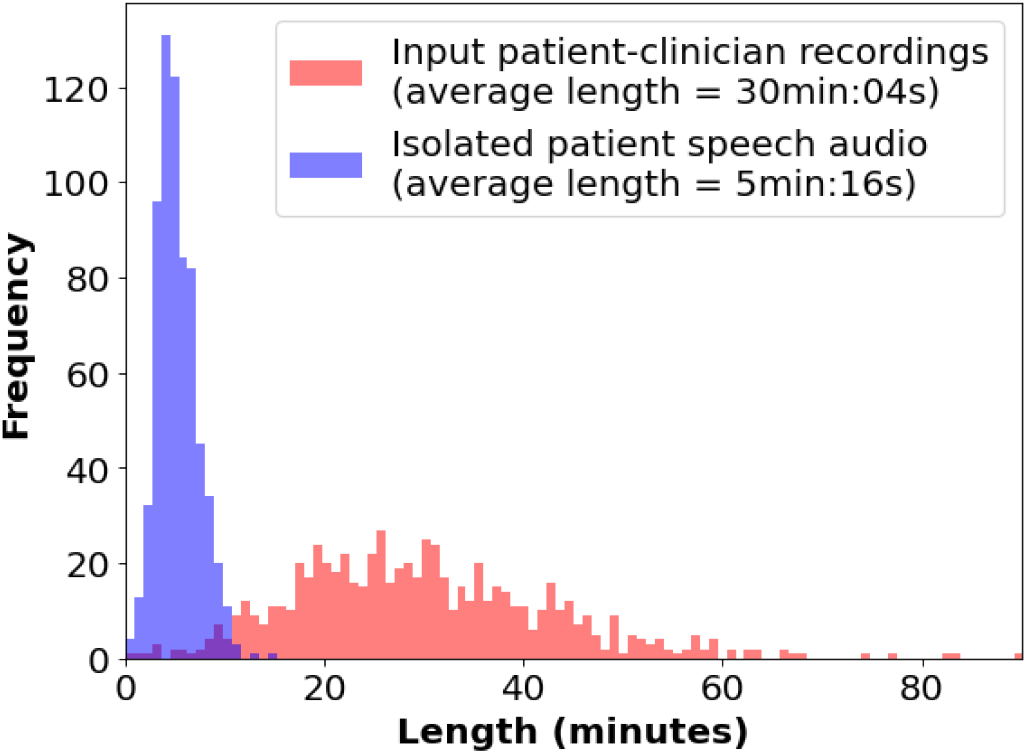
Distributions of audio file lengths for our entire audio dataset, both for the original patient-clinician conversations (red bars) and the output audio of patient speech isolated using our algorithm (blue bars). Our algorithm reduced audio lengths from an average of 30 minutes and 4 seconds to 5 minutes and 15 seconds.

## IV. Conclusions and discussion

In this paper, we proposed an algorithm for identifying and isolating patient speech from unstructured clinical audio, specifically conversations between patients and primary care clinicians. Going beyond the diarization tools used in clinical studies, the proposed solution also separates the audio buffers of different speakers, and identifies which one corresponds to the patient. This enables researchers to access isolated patient speech without having to train or implement specialized diarization tools into their workflow, allowing them to focus on developing models for diagnosing disorders or developing novel disease biomarkers from speech.

The proposed algorithm was evaluated using multiple validation sets sampled from a large and novel dataset of audio recordings of patient-clinician conversations during primary care visits. This evaluation leveraged human experts capable of distinguishing patient speech from other components of the recordings, e.g., clinician and other speech and background noise. Among the multiple implementations of the algorithm tested, the most effective version was able to almost perfectly isolate patient speech segments. We also qualitatively analyzed factors affecting this performance, and illustrated savings of computational resources that could enhance the computational efficiency of downstream analysis algorithms.

Despite the advance for clinical audio processing and ASA represented by our work, it has several limitations that suggest directions for future work. Most prominently, our assessment of the accuracy of the proposed algorithm was limited to short segments of patient speech and relatively small validation sets. This was due to the labor-intensive nature of labeling long patient-clinician conversations in terms of speakers at instances throughout the recording. It would be beneficial to comprehensively quantify the sensitivity of our and other such algorithms by establishing a “gold standard” of recordings labeled in terms of speakers, e.g., patients, clinicians and other speakers. Our algorithm also leveraged the placement of the lavalier microphone on the patient, as is commonplace in clinical research settings [8], [17]. Thus, it’s performance may not translate for other recording settings, which may need their own specialized algorithms. Another limitation was that we adopted heuristic methods for isolating a patient’s speech from other parts of the recording, each of which had strengths and weaknesses, as illustrated in Section III-B. It would be beneficial to systematize these methods and integrate them with established diarization algorithms, such as those in pyannote-audio used in this work. Finally, our algorithm utilized only the audio signal, not the content of the conversation between the patient and clinician. It would be more effective to integrate both these streams of information to improve the diarization and patient speech isolation steps.

## Data Availability

Due to restrictions in the IRB approval and the presence of PHI, the data can not be released publicly.

## Acknowledgment

This work was supported by NIH grant #s R01 AG066471 and R01 HG011407. The technical parts of this work were supported in part by Oracle Cloud credits and related resources provided by the Oracle for Research program, as well as the computational resources and staff expertise provided by Scientific Computing at the Icahn School of Medicine at Mount Sinai. We also thanks Yan Chak Li and Kaleigh Fidaleo for their technical assistance.

## Notes

### Competing Interest Statement

The authors have declared no competing interest.

### Funding Statement

This work was supported by NIH grants R01 AG066471 and R01 HG011407. The technical parts of this work were supported in part by Oracle Cloud credits and related resources provided by the Oracle for Research program, as well as the computational resources and staff expertise provided by Scientific Computing at the Icahn School of Medicine at Mount Sinai.

### Author Declarations

IRB of the Mount Sinai Health System gave ethical approval for this work.

## References

[1] R. Voleti, J. M. Liss, and V. Berisha, “A review of automated speech and language features for assessment of cognitive and thought disorders,” IEEE Journal of Selected Topics in Signal Processing, vol. 14, no. 2, pp. 282–298, 2020.

[2] L. S. Bickley, P. G. Szilagyi, R. M. Hoffman, and R. P. Soriano, Bates’ pocket guide to physical examination and history taking. Lippincott Williams & Wilkins, 2020.

[3] J. Zhang, Z. Pan, C. Gui, T. Xue, Y. Lin, J. Zhu, and D. Cui, “Analysis on speech signal features of manic patients,” Journal of Psychiatric Research, vol. 98, pp. 59–63, 2018.

[4] A. König, A. Satt, A. Sorin, R. Hoory, O. Toledo-Ronen, A. Derreumaux, V. Manera, F. Verhey, P. Aalten, P. H. Robert, and R. David, “Automatic speech analysis for the assessment of patients with predementia and alzheimer’s disease,” Alzheimer’s & Dementia: Diagnosis, Assessment & Disease Monitoring, vol. 1, no. 1, pp. 112–124, 2015.

[5] M. L. B. Pulido, J. B. A. Hernández, M. Á . F. Ballester, C. M. T. González, J. Mekyska, and Z. Smékal, “Alzheimer’s disease and automatic speech analysis: a review,” Expert systems with applications, vol. 150, p. 113213, 2020.

[6] E. Alpaydin, Machine learning. MIT Press, 2021.

[7] I. Goodfellow, Y. Bengio, and A. Courville, Deep learning. MIT press, 2016.

[8] A. Kumar, T. Jaquenoud, J. H. Becker, D. Cho, M. R. Mindt, A. Federman, and G. Pandey, “Can you hear me now? clinical applications of audio recordings,” medRxiv, 2022.

[9] X. Anguera, S. Bozonnet, N. Evans, C. Fredouille, G. Friedland, and O. Vinyals, “Speaker diarization: A review of recent research,” IEEE Transactions on Audio, Speech, and Language Processing, vol. 20, no. 2, pp. 356–370, 2012.

[10] S. Tranter and D. Reynolds, “An overview of automatic speaker diarization systems,” IEEE Transactions on Audio, Speech, and Language Processing, vol. 14, no. 5, pp. 1557–1565, 2006.

[11] J. Weiner, M. Angrick, S. Umesh, and T. Schultz, “Investigating the effect of audio duration on dementia detection using acoustic features,” 09 2018, pp. 2324–2328.

[12] T. Al Hanai, R. Au, and J. Glass, “Role-specific language models for processing recorded neuropsychological exams,” in Proceedings of the 2018 Conference of the North American Chapter of the Association for Computational Linguistics: Human Language Technologies, Volume 2 (Short Papers). New Orleans, Louisiana: Association for Computational Linguistics, Jun. 2018, pp. 746–752.

[13] H. Lin, C. Karjadi, T. F. A. Ang, J. Prajakta, C. McManus, T. W. Alhanai, J. Glass, and R. Au, “Identification of digital voice biomarkers for cognitive health.” in Exploration of medicine vol. 1, Dec. 2020, p. 406–417.

[14] C. Garoufis, A. Zlatintsi, P. P. Filntisis, N. Efthymiou, E. Kalisperakis, Garyfalli, T. Karantinos, L. Mantonakis, N. Smyrnis, and P. Maragos, “An unsupervised learning approach for detecting relapses from spontaneous speech in patients with psychosis,” in 2021 IEEE EMBS International Conference on Biomedical and Health Informatics (BHI), 2021, pp. 1–5.

[15] L. Hansen, Y.-P. Zhang, D. Wolf, K. Sechidis, N. Ladegaard, and R. Fusaroli, “A generalizable speech emotion recognition model reveals depression and remission,” Acta Psychiatrica Scandinavica, vol. 145, no. 2, pp. 186–199, 2022.

[16] H. Bredin, R. Yin, J. M. Coria, G. Gelly, P. Korshunov, M. Lavechin, D. Fustes, H. Titeux, W. Bouaziz, and M.-P. Gill, “pyannote.audio: neural building blocks for speaker diarization,” 2019.

[17] N. Cummins, S. Scherer, J. Krajewski, S. Schnieder, J. Epps, and T. F. Quatieri, “A review of depression and suicide risk assessment using speech analysis,” Speech Communication, vol. 71, pp. 10–49, jul 2015.

[18] N. Ryant, P. Singh, V. Krishnamohan, R. Varma, K. Church, C. Cieri, J. Du, S. Ganapathy, and M. Liberman, “The third dihard diarization challenge,” arXiv preprint arXiv:2012.01477, 2020.

[19] A. V. Oppenheim, J. R. Buck, and R. W. Schafer, Discrete-time signal processing. Vol. 2. Upper Saddle River, NJ: Prentice Hall, 2001.

[20] G. Van Rossum and F. L. Drake, Python 3 Reference Manual. Scotts Valley, CA: CreateSpace, 2009.

[21] A. Paszke, S. Gross, F. Massa, A. Lerer, J. Bradbury, G. Chanan, T. Killeen, Z. Lin, N. Gimelshein, L. Antiga, A. Desmaison, A. Kopf, E. Yang, Z. DeVito, M. Raison, A. Tejani, S. Chilamkurthy, B. Steiner, L. Fang, J. Bai, and S. Chintala, “Pytorch: An imperative style, high-performance deep learning library,” in Advances in Neural Information Processing Systems 32. Curran Associates, Inc., 2019, pp. 8024–8035. [Online]. Available: http://papers.neurips.cc/paper/9015-pytorch-an-imperative-style-high-performance-deep-learning-library.pdf

[22] C. Angermueller, T. Pärnamaa, L. Parts, and O. Stegle, “Deep learning for computational biology,” Molecular systems biology, vol. 12, no. 7, p. 878, 2016.

